# Neuroradiological findings in GAA-*FGF14* ataxia (SCA27B): more than cerebellar atrophy

**DOI:** 10.1101/2024.02.16.24302945

**Authors:** Shihan Chen, Catherine Ashton, Rawan Sakalla, Guillemette Clement, Sophie Planel, Céline Bonnet, Phillipa Lamont, Karthik Kulanthaivelu, Atchayaram Nalini, Henry Houlden, Antoine Duquette, Marie-Josée Dicaire, Pablo Iruzubieta Agudo, Javier Ruiz Martinez, Enrique Marco de Lucas, Rodrigo Sutil Berjon, Jon Infante Ceberio, Elisabetta Indelicato, Sylvia Boesch, Matthis Synofzik, Benjamin Bender, Matt C. Danzi, Stephan Zuchner, David Pellerin, Bernard Brais, Mathilde Renaud, Roberta La Piana

**Author notes:** **Corresponding Author** Roberta La Piana, Department of Neurology & Neurosurgery and Department of Diagnostic Radiology Montreal Neurological Institute and Hospital, McGill University, 3801 rue University, Montreal, Quebec, Canada H3A 2B4.

## Abstract

**Background:** GAA-*FGF14* ataxia (SCA27B) is a recently reported late-onset ataxia caused by a GAA repeat expansion in intron 1 of the *FGF14* gene. Initial studies revealed cerebellar atrophy in 74-97% of patients. A more detailed brain imaging characterization of GAA-*FGF14* ataxia is now needed to provide supportive diagnostic features and earlier disease recognition.

**Methods:** We performed a retrospective review of the brain MRIs of 35 patients (median age at MRI 63 years; range 28-88 years) from Quebec (n=27), Nancy (n=3), Perth (n=3) and Bengaluru (n=2) to assess the presence of atrophy in vermis, cerebellar hemispheres, brainstem, cerebral hemispheres, and corpus callosum, as well as white matter involvement. Following the identification of the superior cerebellar peduncles (SCPs) involvement, we verified its presence in 54 GAA-*FGF14* ataxia patients from four independent cohorts (Tübingen n=29; Donostia n=12; Innsbruck n=7; Cantabria n=6). To assess lobular atrophy, we performed quantitative cerebellar segmentation in 5 affected subjects with available 3D T1-weighted images and matched controls.

**Results:** Cerebellar atrophy was documented in 33 subjects (94.3%). We observed SCP involvement in 22 subjects (62.8%) and confirmed this finding in 30/54 (55.6%) subjects from the validation cohorts. Cerebellar segmentation showed reduced mean volumes of lobules X and IV in the 5 affected individuals.

**Conclusions:** Cerebellar atrophy is a key feature of GAA-*FGF14* ataxia. The frequent SCP involvement observed in different cohorts may facilitate the diagnosis. The predominant involvement of lobule X correlates with the frequently observed downbeat nystagmus.

## Introduction

Late-onset cerebellar ataxias (LOCA), defined by the onset of progressive cerebellar syndrome after 30 years of age, are heterogenous neurodegenerative disorders that have largely resisted molecular diagnosis until recently.^1, 2^ GAA-*FGF14* ataxia (spinocerebellar ataxia SCA27B; OMIM 620174) is a recently described LOCA caused by a dominantly inherited GAA-TCC repeat expansion in the first intron of *FGF14*, which encodes the fibroblast growth factor-14.^3, 4^ This disorder is estimated to be one of the most common forms of late-onset as well as autosomal dominant ataxias, especially in the French Canadian (around 60% of previously undiagnosed patients with LOCA) and European population (around 20% of all autosomal dominant ataxias).^5^ Initial review of brain MRI findings has revealed cerebellar atrophy in 74% of GAA-*FGF14* ataxia patients, with further data reporting vermian atrophy in over 90% of patients.^3, 6^ It is now important to precisely define the neuroradiological features of GAA-*FGF14* ataxia that distinguish it from other LOCAs, to 1) provide supportive diagnostic features to ensure early diagnosis, and 2) explore its pathophysiological bases.

## Methods

We performed a retrospective analysis of MRI images of patients with GAA-*FGF14* ataxia. Subjects were included if a) their phenotype was clinically compatible with GAA-*FGF14* ataxia, b) the genetic testing documented a (GAA) expansion in *FGF14*, according to Bonnet et al.,^7^ and c) available MRI scans included at least sagittal or 3D T1- or 3D FLAIR (Fluid-attenuated inversion recovery) T2-weighted images.^7, 8^ Subjects were recruited in Quebec (n=27), Nancy (n=3), Perth (n=3) and Bengaluru (n=2). For all subjects, we collected demographic and clinical data.

All images were reviewed collegially by the authors (SC, RS, CA, and RLP) according to the qualitative approach detailed below and blind to disease severity and presented symptoms. Each MRI scan was evaluated twice, 3 months apart. Team disagreements were resolved by consensus. The institutional review boards of each participating institution approved the use of clinical data for the retrospective study.

### Qualitative MRI analysis

We performed a qualitative analysis to assess the degree of atrophy in the vermis, cerebellar hemispheres, and cerebral hemispheres (size of ventricular system and CSF spaces). For the vermis the atrophy was assessed on midline sagittal planes as ‘present’ or ‘absent’, with further grading as ‘mild’, ‘moderate’ or ‘severe’. For the cerebellar and cerebral hemispheres, the atrophy was assessed on all planes and, when present, graded as ‘mild’, ‘moderate’ or ‘severe’. The effect of the patient’s age was taken into consideration by comparing the images to standard image series used in a study of a neurologically healthy population.^9^

To assess the presence of brainstem atrophy we measured the maximal anteroposterior diameter at the level of the midbrain, midpons, and medulla on sagittal images, and compared to normative values for age according to Metwally et al.^10^ The corpus callosum thickness was quantified by measuring the maximum width of the body at midpoint and compared to normative values according to Gupta et al.^11^

We also assessed the presence and characteristics of white matter abnormalities on T2- and FLAIR T2-weighted images on all planes, when available, by location (periventricular, subcortical, juxtacortical), lobar involvement (frontal, parietal, temporal, occipital, cerebellar, brainstem), and symmetry. Juxtacortical lesions were defined as lesions in direct contact with the cortical ribbon. Periventricular lesions included all lesions abutting any portion of lateral ventricles. Subcortical lesions were defined as supratentorial non-juxtacortical and non-periventricular. As for the assessment of atrophy described above, the effect of the patient’s age was taken into consideration by comparing to standard images and prevalence across age from previous studies of healthy subjects.^9, 12^

The protocols of the reviewed MRIs were not standardized, as they were requested for clinical indication across different hospitals and scanners, thus limiting the systematic analysis of MR data.

### Clinical-radiological correlations

We compared the prevalence of radiological findings (cerebral, cerebellar and vermian atrophy; superior cerebellar peduncle (SCP) involvement, periventricular and multifocal white matter abnormalities; corpus callosum thinning; ventricular enlargement) between patients symptomatic for ≤ vs. > than 5 years (n=11 and n=24, respectively) at the time of initial MRI scan, since most patients become permanently ataxic after an average of 5 years since disease onset, and between patients aged ≤ vs > 60 years-old (n=14 and n=21 respectively) at the time of their initial MRI. We assessed differences between groups with the Fisher’s exact test. Additionally, we sought to compare the radiological findings between subjects with episodic ataxia vs. permanent ataxia at the time of MRI, and between subjects with GAA expansion size <300 vs. ≥300. However, the highly unequal sample size between groups prevented optimal statistical testing. Results from all analyses are included in the supplementary material (**Supplementary Table 3**).

#### Secondary Analysis

Following the identification of the involvement of the SCPs, we sought to verify its presence in four independent cohorts from Tübingen (Germany), Innsbruck (Austria), Santander (Spain), and San Sebastian (Spain). Neurologists and radiologists from each center (M.S. and B.Be., University of Tübingen; E.I., S. B. University of Innsbruck; P.I.A, A.L.M.A., University of Donostia; J.I.C., E.M.L., R.S.B. University of Cantabria) reviewed the brain MR images of their GAA-*FGF14* ataxia patients (Tübingen n=31; Donostia n=14; Innsbruck n=9; Cantabria n=6) to assess the presence of the SCP involvement comparing to provided reference images (**Supplementary Figure 1**). The Montreal group also independently reviewed the same MRIs, blind to the treating teams’ conclusions. Disagreement between the two assessments were solved by consensus.

### Measurements and Quantitative MRI analysis

To assess the degree of cerebellar atrophy, we performed quantitative MRI analyses on individuals with confirmed GAA-*FGF14* ataxia *<u>and</u>* available 3D T1-weighted images. Five subjects of the entire cohort (5/35; 14.3%) fulfilled the required imaging criteria. We also performed the quantitative MRI analysis on 5 age-matched healthy controls with available 3D T1-weighted images.

Segmentation of cerebellum using T1-weighted images was performed automatically using the CERES pipeline (volBrain)^13^ (**Supplementary Figure 2**). All anatomical structures were defined based on the MRI atlases described in Park et al., 2014.^14^ We compared the following parameters between affected individuals and controls: total and lobular cerebellum volume (grey matter and white matter), cerebellar cortical thickness, and asymmetry index. Cerebellar volume percentage was calculated as the ratio between cerebellar volume and total intracranial volume.

## Results

We reviewed brain MRI studies of 35 patients (14 not previously reported): 15 females, median age at MRI was 63 years (range 28-88 years); median disease duration at the time of MRI was 9 years (range 1-34 years), median age of disease onset at 52 years (range 25-75 years). The clinical and radiological features of the cohort are summarized in **Table 1**. Longitudinal studies were also available for seven subjects. In terms of disease duration, 24 patients (54.3%) were symptomatic for >5 years at the time of initial MRI.

**Table 1.** Demographic, clinical and radiological findings of 35 subjects with GAA-*FGF14* ataxia. *Legend*: SCP=superior cerebellar peduncle involvement; M = multifocal white matter abnormalities; PB = Periventricular, around anterior and posterior horns; PA = Periventricular, around anterior horns only; PP= Periventricular, around posterior horns only. *Data missing for 4 subjects.

| **Demographics and clinical information** | **n (%) - tot 35** |
| --- | --- |
| Sex F;M | 15; 20 (42.8; 57.2) |
| Median age at onset, years [range] | 52 [25-75] |
| Median age at MRI, years [range] | 63 [28-88] |
| Median disease duration, years [range] | 9 [1-34] |
| Median GAA-TCC repeat expansion [range] | 378 [265-508] |
| Permanent ataxia at time of MRI* | 29 (93.5) |
| Episodic ataxia at time of MRI* | 2 (6.5) |
| Down beat nystagmus | 20 (57.1) |
| Gaze-evoked horizontal nystagmus | 20 (57.1) |
| Vascular risk factors | 19 (54.3) |
| **MRI findings** | **n (%); tot 35** |
| *Cerebral atrophy* | *15 (42.8)* |
| mild | 11 (31.4) |
| moderate | 4 (11.4) |
| *Cerebellar involvement* | *33 (94.3)* |
| *Vermian atrophy* | *33 (94.3)* |
| mild | 13 (37.1) |
| moderate | 17 (48.6) |
| severe | 3 (8.6) |
| *Brainstem atrophy* | *1 (2.8)* |
| *Hemispheric atrophy* | *22 (62.8)* |
| mild | 15 (42.8) |
| moderate | 6 (17.1) |
| severe | 1 (2.8) |
| *White matter abnormalities* | *34 (97.1)* |
| SCP | 22 (62.8) |
| multifocal | 22 (62.8) |
| periventricular | 28 (80) |
| *Corpus callosum thinning* | *7 (20)* |
| *Ventricular enlargement* | *13 (37.1)* |
| 4<sup>th</sup> ventricle | 4 (11) |
| Extended to lateral ventricles | 9 (25.7) |

### Radiological Features

#### MRI patterns of atrophy

##### Infratentorial atrophy

Cerebellar atrophy was observed in 33 patients (33/35, 94.3%), including 22 patients (22/35, 62.8%) with involvement of both vermis and cerebellar hemispheres, and 11 (11/35, 31.4%) with isolated vermian atrophy **(Figure 1**).

**Figure 1.**
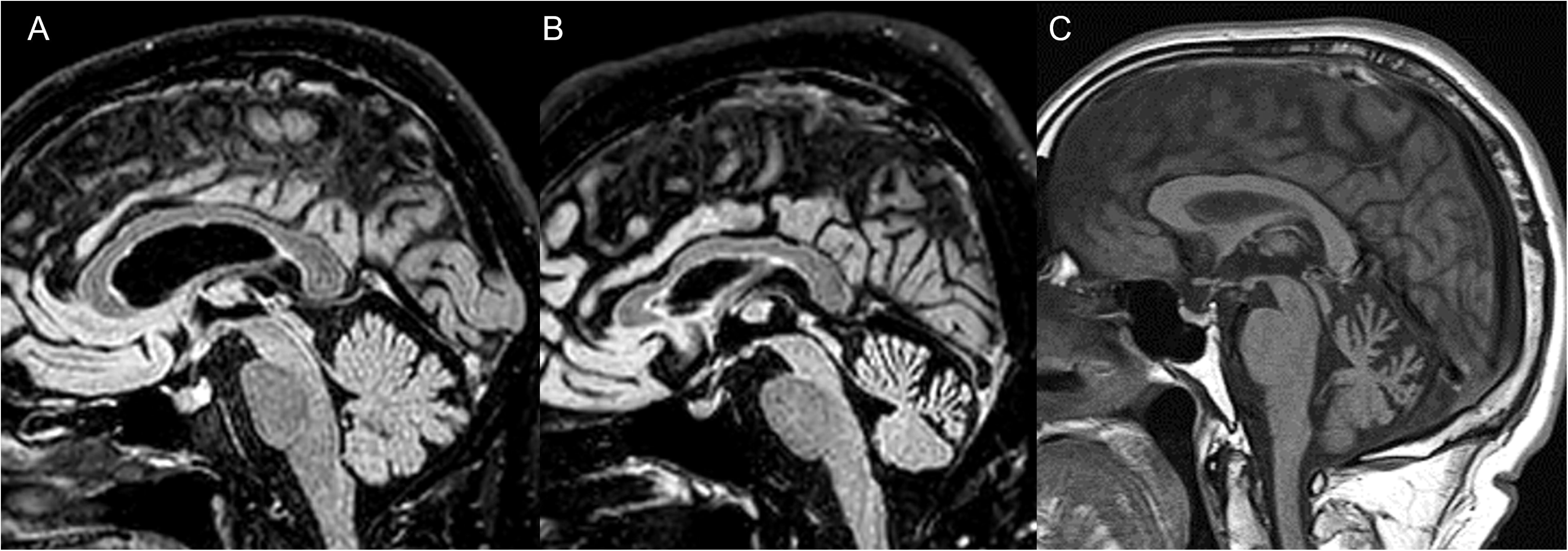
Cerebellar atrophy. Sagittal Flair T2 (A, B) and T1-weighted images of three subjects showing different degrees of cerebellar atrophy. A, subject in their 60s with 9 years of disease duration with normal size of vermis (GAA-TCC expansion size 374); B, moderate cerebellar atrophy of the anterior and superior lobe of the vermis in a subject in their 60s with 34 years of disease duration (GAA-TCC expansion size 374); C, severe cerebellar atrophy of the anterior and superior lobe of the vermis in a subject in their 60s with 14 years of disease duration (GAA-TCC expansion size 437).

For the 33 patients with vermian atrophy – either isolated or not – the degree was mild in 13 (13/35, 37.1%), moderate in 17 (17/35, 48.5%), and severe in 3 (3/35, 8.5%). The majority of patients (32/35, 91.4%) had predominant involvement of the anterior lobe and the superior posterior lobe of the vermis before the pre-pyramidal fissure, except for one individual with diffuse atrophy involving the vermis globally.

Among the 22 patients with both vermian and hemispheric involvement, cerebellar hemispheres were mildly atrophic in 15 subjects (15/35, 42.8%), moderately atrophic in six (6/35, 17.1%), and severely atrophic in one (1/35, 2.9%). The degree of atrophy was the same in both the vermis and cerebellar hemispheres in 11 (50%), relatively more severe in the vermis compared to the hemispheres in ten (10/22, 45.5%) and more severe in the cerebellar hemispheres in one patient (4.5%).

We documented reduced brainstem anteroposterior diameter in one subject (1/35; 2.8%) (AP diameter 1.77mm, normal range for age 1.9-2.5) who presented concomitant vermian and cerebellar hemispheric atrophy as well as SCP involvement.

##### Supratentorial atrophy

Cerebral atrophy was present in 15 subjects (15/35, 42.9%; median age 69 years, range 45-80): mild in 11 (11/35, 31.4%; median age 63 years, range 45-80), and moderate in four (4/35, 11.4%; median age 73 years, range 67-77). This was particularly evident in terms of increased CSF spaces in the frontal lobes.

Ventricular enlargement was documented in 13 subjects (13/35, 37.1%); limited to the 4^th^ ventricle in four (4/35, 11.4%) and extended to the lateral ventricles in 9 (9/35, 25.7%).

We documented reduced corpus callosum body thickness for age in 7 subjects (7/35; 20%): (0.32, 0.35, 0.38, 0.39, 0.44, 0.48, 0.48mm; mean value for age 0.65 ± 0.15).

##### Longitudinal studies

The mean interval between MRIs was 6.4 years (range 1-16 years). All seven patients had cerebellar atrophy on the initial MRI, which remained stable in two patients and progressed in five (5/7, 71.4%) (**Figure 2**). We also observed progression of cerebral atrophy in one patient, likely related to age (1/7, 14.3%; age at MRI 63 and 79 years) (**Figure 2D**) and progression of ventricular enlargement in two (2/7, 28.6%). Two patients (2/7, 28.6%) developed corpus callosum thinning on subsequent MRI (**Figure 2D**).

**Figure 2.**
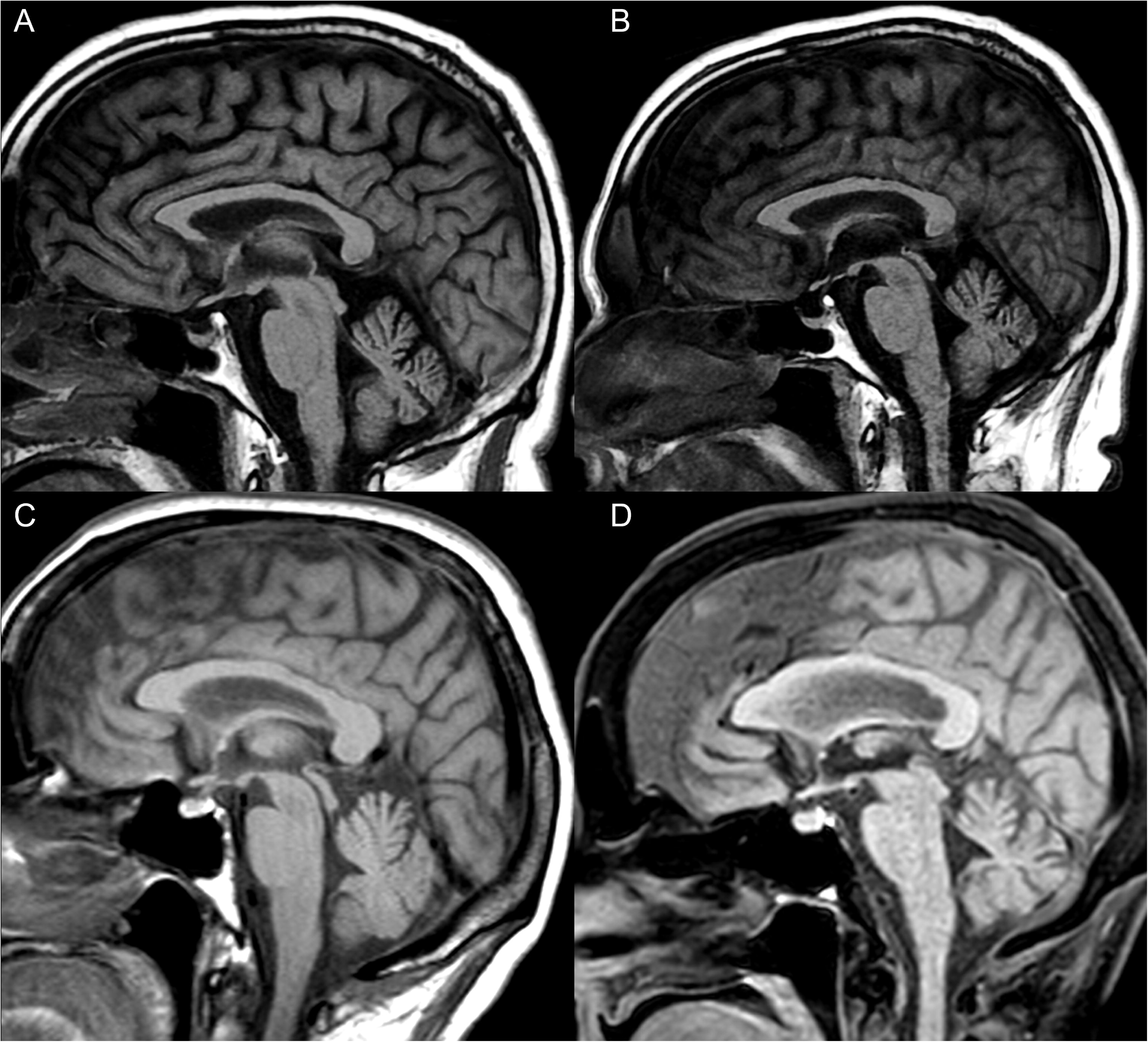
Cerebellar atrophy, longitudinal data. A,B. Sagittal T1-weighted images of a patient (GAA-TCC expansion size 424) showing stable degree of mild cerebellar atrophy localized mainly in the anterior lobe of the vermis, over the course of 10 years, from their 40s, 17 years after disease onset (A), to their 50s (B). C,D. Sagittal T1-weighted images of a patient showing progression of the vermian atrophy that extended from the anterior lobe of the vermis (C, while in their 60s, 5 years after disease onset) to the posterior aspect of the vermis (D), 16 years after the first MRI. In addition, the images show that the patient developed mild corpus callosum thinning at the level of the body and midline cerebral atrophy over time.

#### MRI patterns of white matter abnormality

T2 or FLAIR T2-weighted images on all three planes were available for 19 subjects, while for the remaining 16 only axial T2 or FLAIR T2 images were available for review.

White matter abnormalities (WMAs) were present in 34 subjects (34/35, 97.1%; median age 63, range 28-88). Different patterns were observed: a characteristic SCP involvement (**Figure 3**), as well as non-specific periventricular and multifocal abnormalities (**Figure 4**).

**Figure 3.**
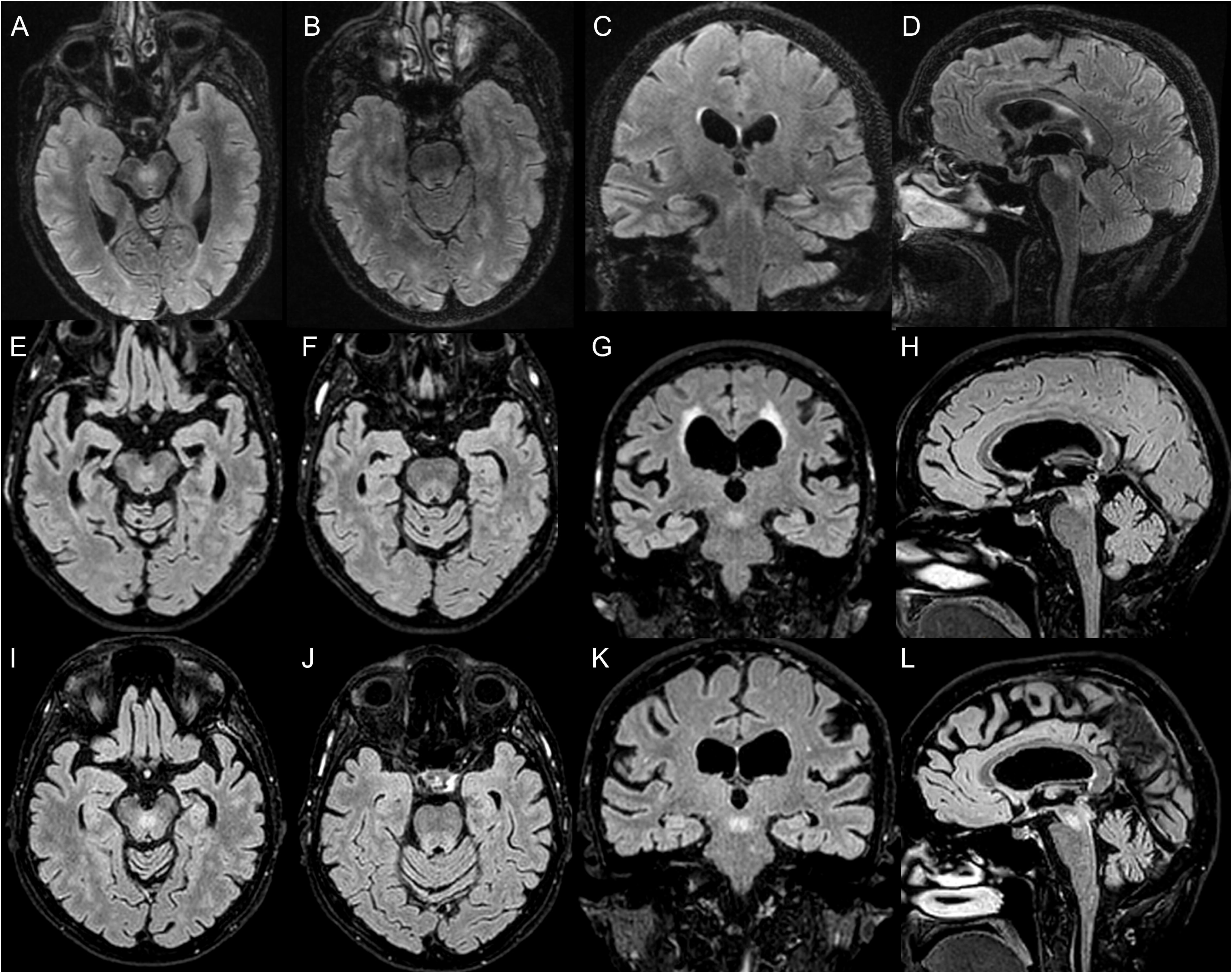
Involvement of the superior cerebellar peduncles. Multiplanar Flair T2-weighted images of three subjects showing bilateral and symmetric involvement of the superior cerebellar peduncles and its decussation within the midbrain. T2-hyperintense signal is visible in the central region of the midbrain both in the axial (A, E, I) and sagittal views (D, H, L), as well as in the coronal plane (C, G, K). The SCPs are involved for their entire length, as showed in the axial views (B, F, J) and in the coronal image (C). The images are from a subject in their 70s with 5 history of disease duration (GAA-TCC expansion size 324) (A-D), a subject in their 80s with 10 years of disease duration (E-H) (GAA-TCC expansion size 250), a subject in their 70s with 21 years of disease duration (GAA-TCC expansion size 357) (I-L).

**Figure 4.**
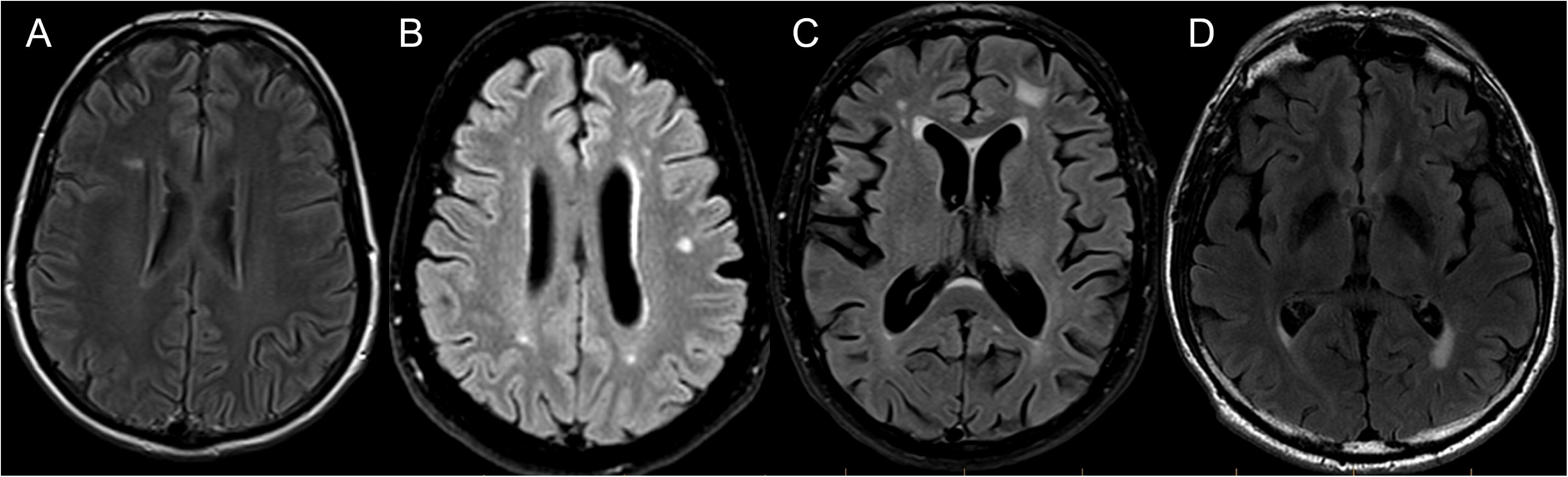
Non-specific white matter abnormalities. Axial FLAIR T2-weighted images showing different supratentorial white matter abnormalities. A, a focal oval-shaped lesion in the right frontal deep white matter in a subject in their 70s (GAA-TCC expansion size 357); B, multifocal subcortical white matter abnormalities in a subject in their 70s (GAA-TCC expansion size 330); C, symmetric periventricular white matter involvement with enlargement of the lateral ventricles and multifocal subcortical white matter abnormalities in a subject in their 50s (GAA-TCC expansion size 304); D, posterior periventricular white matter involvement in a subject in their 70s (GAA-TCC expansion size 290). None of these subjects were known for vascular risk factors.

##### SCP involvement

We observed involvement of the SCP and their decussation within the midbrain (**Figure 3**) in 22 subjects (22/35, 62.8%), 16 (16/22, 72.7%) of which had prominent hyperintensity (**Figure 3A-D, I-L**), while six (6/22, 27.2%) presented with more faint changes (**Figure 3E-H**). SCP changes were best captured by 3D FLAIR T2 images and prominent hyperintensity was visible in all three planes (coronal, sagittal, and axial), when available.

###### SCP involvement in validation cohorts

From the 60 subjects with confirmed GAA-*FGF14* ataxia from the four independent cohorts, we reviewed T2-weighted and FLAIR T2-weighted images of 54 subjects (Tübingen = 29; Donostia = 12; Innsbruck = 7; Cantabria = 6). Six subjects were excluded for poor image quality. Involvement of the SCP was identified in 30 subjects (30/54, 55.6%). Fifteen of them (15/30; 50%) had prominent T2 or FLAIR T2 hyperintense signal along the SCP, while 15 (15/30; 50%) had faint signal change. **Supplementary Table 1** includes the results for each independent cohort.

##### Non-specific periventricular and multifocal WMA

Periventricular WMAs were present in 28 (28/35, 80%; median age 64, range 28-88) cases: in six (6/35, 17.1%) they were limited to the white matter adjacent to the anterior horns of the lateral ventricles, in seven to the posterior horns only (7/35, 20%), and in 14 they extended to both anterior and posterior horns (15/35, 42.9%) (**Figure 4C, D**). A minority (6/35, 17.1%) had more diffuse involvement with extension to the peritrigonal area (**Figure 4B**). Non-specific subcortical multifocal lesions were observed in 22 subjects (22/35, 62.8%; median age 64, range 44-88) (**Figure 4A, B, C**). **Supplementary Table 2** includes the prevalence and p values of periventricular and multifocal abnormalities in the total sample and in different age groups as compared to a previously reported healthy cohort.^12^ The prevalence of periventricular white matter abnormalities was significantly higher (p<0.05) in subjects with *GAA-FGF14* ataxia when compared to the healthy population cohort.

Most patients (29/35, 82.9%) had more than one type of WMA: 12 presented periventricular, multifocal, and SCP involvement (12/35, 34.3%), seven (7/35, 20%) had periventricular and multifocal lesions, six (6/35, 17.1%) had both SCP involvement and periventricular changes, and the four (4/35, 11.4%) had SCP involvement and multifocal abnormalities.

Vascular risk factors were documented in 19 patients (19/35, 54.3%). Among these, six (6/19, 31.5%) presented with SCP abnormal signal, ten (10/19, 52.6%) with periventricular white matter involvement, ten (10/19, 52.6%) with multifocal lesions, and seven (7/19, 36.8%) with ventricular enlargement. Approximately one third (6/19, 31.5%) presented with both non-specific white matter lesions and ventricular enlargement.

##### Longitudinal studies

Among the seven patients with longitudinal MRI studies, SCP involvement was observed on the initial scan in five (5/7, 71.4%), and remained stable overtime. One case showed new onset mild SCP involvement 4 years after the initial MRI study. Progression of periventricular and multifocal white matter abnormalities was noted in one (1/7, 14.3%) and four (4/7, 57.1%) patients, respectively.

#### Clinical-radiological correlations

We did not document any statistically significant differences in the presence of neuroradiological findings between patients with ≤vs. > 5 years of disease duration. Cerebral atrophy and ventricular enlargement were more prevalent in older patients (>60 years-old at initial MRI) compared to the rest of the cohort (p=0.046; p=0.033, respectively). Conversely, isolated vermian atrophy and involvement of the SCP were more prevalent in younger patients (≤60 years-old at MRI) (p=0.023 and p<0.001, respectively). All the *p* values relative to the comparison between groups are reported in **Supplementary Table 3**.

#### Quantitative analysis of cerebellar atrophy

##### Cerebellar Volume

**Table 2** includes the results of the quantitative cerebellar volume analysis for the total volume percentage as well as grey matter percentage for the entire cerebellum and individual lobules in the five patients with available 3D T1-weighted images and controls.

**Table 2.**
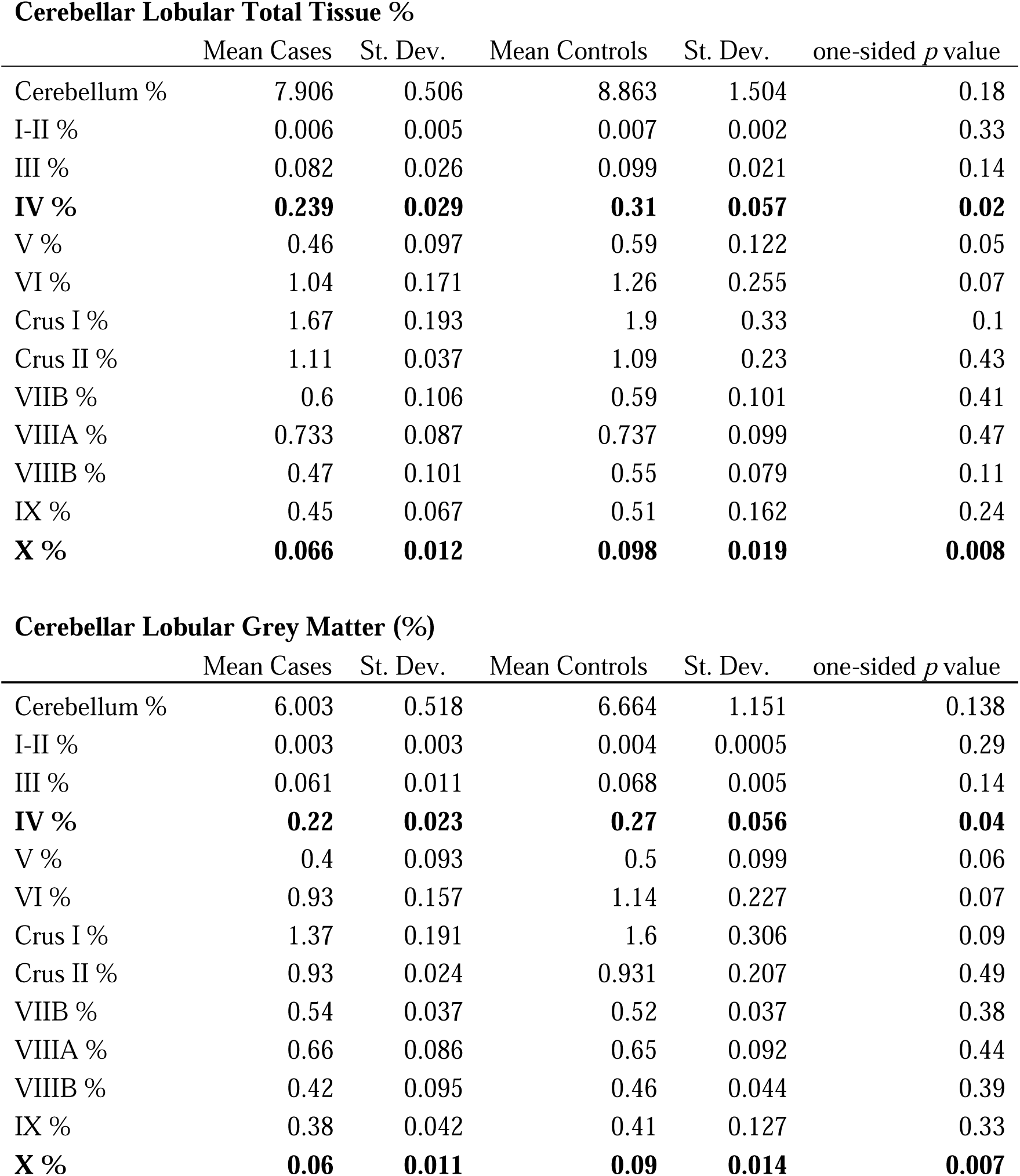
Quantitative cerebellar volume analysis in 5 subjects with GAA-*FGF14* ataxia and available high quality sagittal or 3D T1-weighted images and age-matched controls. Mean values, standard deviations, and one-sided p values for the individual lobules for patients (cases) and controls. In bold the statistically significant differences (p<0.05) between groups.

We did not observe significant differences in the total cerebellar volume nor total grey matter volume between patients with GAA-*FGF14* ataxia and controls. Analyses of individual lobules revealed a decrease in the total volume percentage of lobule IV (mean 0.24±0.03% vs 0.31±0.06%, t (8) = −2.55, p=0.017, 95%CI [-0.14, −0.007]) and lobule X (mean 0.07±0.01% vs 0.1±0.02%, t (8) = 0.03, p=0.008, 95%CI [-0.056, −0.007]) in patients compared to controls. Analysis of grey matter revealed significant decrease in total volume percentage of lobule IV (mean 0.22±0.02% vs 0.27±0.06, t (8) = −1.99, p=0.04, 95%CI [-0.11, −0.009] and lobule X (mean 0.06±0.01% vs 0.09±0.01%, t (8) =-3.17, p=0.007, 95%CI [-0.046, −0.007]).

Among all lobes, we observed the greatest total volume percentage difference in lobule X (47.4%). In terms of grey matter volume, lobule X showed the greatest reduction in volume percentage (42.5%).

## Discussion

This study confirms that cerebellar atrophy is a common MRI finding in GAA-*FGF14* ataxia and it provides a detailed characterization of it as well as highlights additional neuroradiological features that may facilitate diagnosis.^3, 6, 15^

Cerebellar atrophy of varying severity was observed in 94% of our subjects, all with vermian involvement, and over half with atrophy extending to the cerebellar hemispheres. Vermian atrophy affected predominantly the anterior lobe and the superior part of the posterior lobe (i.e. declive, folium and tuber lobules). Despite the limited number of available longitudinal MR exams, our study documented progression of cerebellar atrophy in more than half of the subjects with available follow up over a median 6.4-year period, in line with a previous study by Wilke et al.^6^

Cerebellar atrophy is the primary radiological finding in patients with cerebellar ataxia.^1, 6, 15^ In LOCA, cerebellar atrophy has been reported in Spinocerebellar Ataxias (SCA) 8 and 17, *RFC1* related disorder and MSA-C (Multiple System Atrophy-Cerebellar type). ^16–22^ Establishing the pattern of atrophy can guide genetic testing and eventual diagnosis. The preferential involvement of the vermis and, to a lesser degree, cerebellar hemispheres can differentiate GAA-*FGF14* ataxia from other LOCA with more widespread atrophy, but the distinction from other LOCAs with more isolated involvement of the cerebellum, such as SCA6, can be more challenging.^18,16^ To attempt to address this, we performed quantitative segmentation of the cerebellum in subjects with available 3D T1-weighted images, which unfortunately were only 14.7% of our cohort, thus limiting the generalizability of our results and needing further confirmation in future prospective studies. In the subjects with GAA-*FGF14* ataxia we assessed, lobules IV and X were preferentially affected, both in total and grey matter volume, with lobule X showing the greatest reduction. Damage to lobule X and flocculus is associated with downbeat nystagmus as hypofunction of lobule X can lead to disinhibition of the superior vestibular nuclei neurons, resulting in spontaneous slow upward drift followed by corrective downward saccade.^23,24^ Gaze-evoked nystagmus is another feature thought to be due to inadequate lobule X control on the brainstem ocular motor integrator.^24^ These results correspond well with 57% of our cohort having downbeat nystagmus, and 57% having gaze-evoked horizontal nystagmus – rates similar to other reported cohorts (range between 37 and 78%).^3, 15, 25, 26^ Nystagmus was present in all the subjects included in the volumetric analysis. Interestingly, *FGF14* (GAA)_≥250_ expansions were documented in 48% of 170 subjects with ‘idiopathic’ downbeat nystagmus.^27^ As proposed by Pellerin et al.^27^, the earliest damage in SCA27B may arise in the flocculus / paraflocculus, thus accounting for the frequently observed early downbeat nystagmus, impaired VOR, other cerebellar ocular motor signs and vertigo. Our data, although preliminary, further support that GAA-*FGF14* ataxia pathology may arise and is particularly severe in this region of the cerebellum. Lobular analysis may aid in the differentiation between LOCAs with similar patterns of global atrophy and should be the focus of future targeted studies. Identification of subregions preferentially affected during the early phase of the disease may promote the development of imaging-markers for early disease detection. As previously found, there is preliminary evidence that some patients with GAA-*FGF14* disease present symptoms limited to the involvement of the cerebellar flocculus / parafloccolus region and do not develop ataxia even after several years of disease duration.^27^

Our study is the first to describe the involvement of the SCPs and their decussation within the midbrain in subjects with GAA-*FGF14* ataxia. While microstructural changes and reduced volume have been documented in SCA2 and Friedreich ataxia respectively,^31–34^ a pattern of abnormal T2-hypersignal along the SCP like the one we observed in GAA-*FGF14* ataxia subjects has been previously documented in some patients with POLR3-related spastic ataxia.^35, 36^ Co-existent SCP and vermian atrophy was present in 57% of our patients, and could suggest that the SCP abnormality is a reflection of reduced vermis-dentate connectivity secondary to vermian atrophy. SCP changes were best captured by 3D T2 FLAIR images. Given the retrospective nature of our study - which limited the availability of ideal sequences - and the presence of suggestive symptoms in GAA-*FGF14* ataxia, it is reasonable to suspect that SCP involvement may be even more prevalent than what we observed.^3, 6^ Hence, we recommend including 3D FLAIR T2-weighted MR sequence when assessing subjects with suspected SCA27B.

Within the limitations of this study, we did not observe any association between disease duration (less vs. more than 5 years) and neuroradiological findings. Cerebral atrophy and ventricular enlargement were more prevalent in older patients. Since these findings can be part of normal aging process or due to other neurodegenerative processes, it will be important to verify these findings in future case-control prospective studies. More interestingly, SCP involvement and vermian atrophy were more represented in patients younger than 60. While it is expected that cerebellar atrophy may be limited to the vermis in younger patients and then extend to cerebellar hemispheres over time, the result is more surprising for the SCP involvement. Several confounding factors might have contributed to this. MRIs included in our study have been acquired over a 20-year span of time and the SCP region may not have been well visualised due to outdated MRI sequences and protocols. Additionally, factors such as family history and access to healthcare resources may affect the timing of one’s initial MRI scan, thus limiting the assessment of differences between groups. Although we cannot completely exclude that the signal abnormalities along the SCP might be transitory, the fact that we documented it in longitudinal exams argues against it. Only prospective dedicated neuroimaging studies will elucidate the diagnostic value of the SCP signal abnormalities, their association with age or clinical features, and their role in the disease pathophysiology. Although formal statistical analysis was prevented by unequal sample sizes between groups, we observed SCP involvement in subjects with episodic ataxia as well as permanent ataxia, and in subjects with (GAA) repeat expansion size below as well as above 300. Therefore, SCP involvement may be a useful additional diagnostic feature, particularly in patients with an incompletely penetrant (GAA)_250-299_ repeat expansion, in earlier stages of disease when only episodic features are observed, and as a marker distinguishing this disorder from other forms of LOCA.^37, 38^

In addition to the SCP involvement, we documented non-specific periventricular and multifocal white matter abnormalities in 80% and over 60% of individuals respectively. As compared to the data from a large healthy population study, the presence of periventricular changes was significantly higher in the total of GAA-*FGF14* ataxia subjects, even though we did not observe statistically significant differences when we performed comparison between age subgroups. The small sample size of the subgroups could contribute to the lack of observed differences with the general population, nonetheless age, cardiovascular risk factors and other neurodegenerative disorders might influence the observed changes.^6, 39^

In conclusion, we described a novel MRI finding of SCP involvement in GAA-*FGF14* ataxia patients, which may be specific to this LOCA subtype and whose detection can orient and accelerate the diagnosis, especially in the early stages of the disease. Our study confirmed that cerebellar atrophy is a key feature of GAA-*FGF14* ataxia, starting at the level of the vermis and extending to the hemispheres over time. The preferential involvement of lobule X correlates with cerebellar ocular motor signs frequently observed in GAA-*FGF14* patients. Lobular volumetric analysis of GAA-*FGF14* ataxia and further dedicated imaging studies may promote diagnostic accuracy, give insights into disease pathogenesis, and contribute to the development of early disease biomarkers.

## Data Availability

All data produced in the present study are available upon reasonable request to the authors.

## Acknowledgements

A. D. Pellerin has received a fellowship award from the Canadian Institutes of Health Research (CIHR). B. Brais has received funds from the Fondation Groupe Monaco and the Canadian Institutes of Health Research (grant 189963). M.C. Danzi and S. Zuckner were supported by the NIH National Institutes of Neurological Disorders and Stroke (grant 2R01NS072248-11A1 to S.Z.) and the NIH National Human Genome Research Institute (grant R21HG013397). M.Synofzik and B. Brais were supported by the European Joint Programme on Rare Diseases, as part of the PROSPAX consortium, under the EJP RD COFUND-EJP N° 825575 (DFG, German Research Foundation, No 441409627). R. La Piana has received a Research Scholar Junior 1 award from the Fonds de Recherche du Québec en Santé (FRQS), and research funds from the Canadian Radiological Foundation, Canadian Institutes of Health Research (grant 506913), Ataxia Canada, the Spastic Paraplegia Foundation and Roche Canada. The authors thank the patients and their families for participating in this study.

## Conflicts of Interests

A. Duquette has received consultancy honoraria from AavantiBio, Novartis, Pfizer Canada, PTC Therapeutics, and Reata Pharmaceuticals, all unrelated to the present manuscript. M. Synofzik has received consultancy honoraria from Ionis, UCB, Prevail, Orphazyme, Servier, Reata, GenOrph, AviadoBio, Biohaven, Zevra, Lilly, and Solaxa, all unrelated to the present manuscript. B. Bender is Co-Founder, shareholder and CTO of AIRAmed GmbH. R. La Piana has received speaking honoraria from Novartis unrelated to the present manuscript.

## Notes

### Author Declarations

The ethics committee of the Montreal Neurological Hospital-Institute gave ethical approval for this work.

### Summary of Updates

Added comparison and statistical analysis for prevalence of white matter abnormalities between GAA-FGF14 ataxia cohort and healthy population study. Discussion modified accordingly.

