## Supplementary Material for "Neuroradiological findings in GAA-*FGF14* ataxia (SCA27B): more than cerebellar atrophy"

**Suppl Figure 1. Assessment of SCP involvement.** The presence of SCP involvement was assessed on T2-weighted images in all three planes, when available. It was graded as faint or prominent according to the intensity of the abnormal signal.

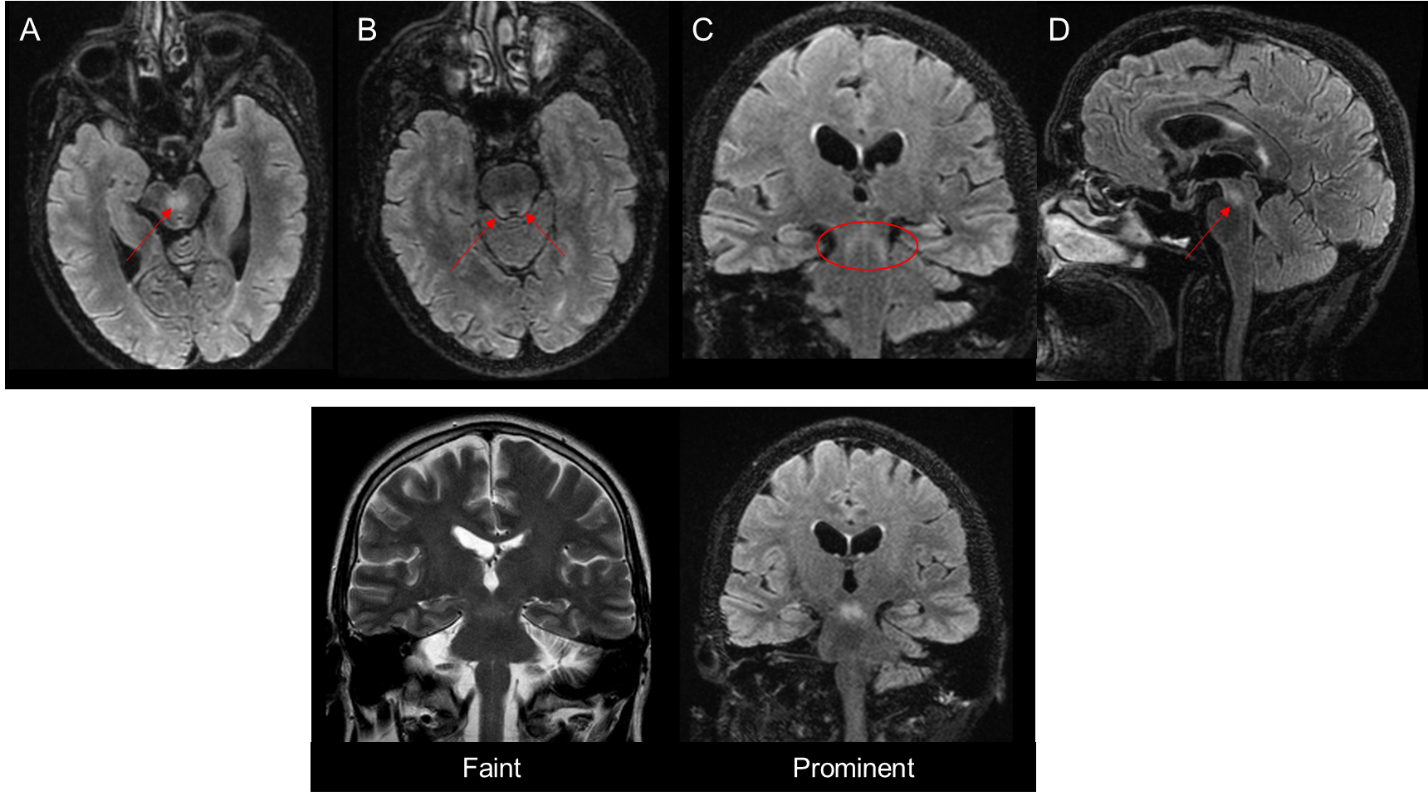

**Suppl Figure 2. CERES cerebellum segmentation.** The following parameters have been computed using the CERES pipeline: total and lobular cerebellum volumes (gray matter and white matter), cerebellar cortical thickness, and hemispherical asymmetry index. Snapshot of CERES labeling steps are included as a quality control.

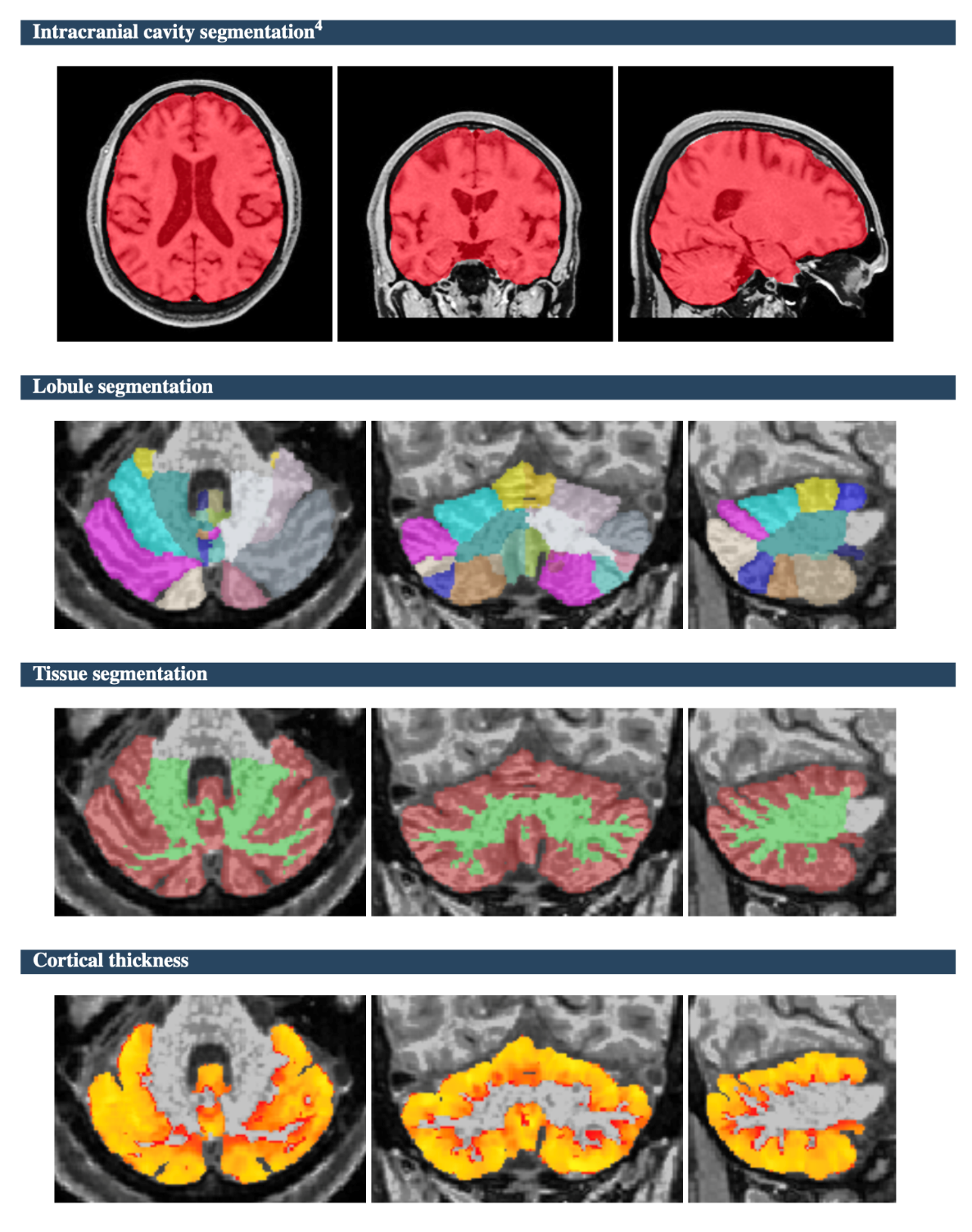

**Supplementary Table 1.** Presence and signal intensity of the SCP involvement in the four independent cohorts used for validation. Percentages refers to each cohort.

| **SCP involvement** | **Total validation cohorts (n=54)** | **Tubingen (n=29)** | **Donostia (n=12)** | **Innsbruck (n=7)** | **Cantabria (n=6)** |
| --- | --- | --- | --- | --- | --- |
| **present** | **30 (55.6%)** | **16 (55.2%)** | **7 (58.3%)** | **4 (57.1%)** | **3 (50%)** |
| prominent | 15 (50%) | 4 (25%) | 7 (100%) | 2 (50%) | 2 (66.7%) |
| faint | 15 (50%) | 12 (75%) | 0 | 2 (50%) | 1 (33.3%) |

**Supplementary Table 2. Periventricular and multifocal white matter abnormalities.** Prevalence and p values in different age groups of GAA-FGF14 ataxia patients as compared to prevalence in healthy subjects from the Austrian Stroke Prevention Study (*Schmidt et al., Acta Neuropathol 2011*). Chi-squared test and Fisher’s exact test were used to assess differences respectively between the total cohorts and the different age groups.

|  | **Periventricular** | | | **Multifocal** |  |  |
| --- | --- | --- | --- | --- | --- | --- |
| **Age group** | **GAA-FGF14 ataxia**  **(N=28)**  n (%) | **Austrian Stroke Prevention Study (N=632)**  n (%) | ***p*** | **GAA-FGF14 ataxia**  **(N=22)**  n (%) | **Austrian Stroke Prevention Study (N=586)**  n (%) | ***p*** |
| Total (GAA-FGF14 N=35; Austrian Study N=993) | 28 | 632 | **0.047** | 22 | 586 | 0.649 |
| ≤54  (GAA-FGF14 N=6; Austrian Study N=128) | 5 (83.3%) | 66 (51.5%) | 0.213 | 4 (66.7%) | 76 (59.3%) | 1 |
| 55-64  (GAA-FGF14 N=14; Austrian Study N=344) | 10 (71.4%) | 203 (59.0%) | 0.417 | 10 (71.4%) | 236 (68.6%) | 1 |
| 65-74  (GAA-FGF14 N=8; Austrian Study N=374) | 6 (75.0%) | 254 (67.9%) | 1 | 4 (50.0%) | 209 (55.8%) | 0.563 |
| ≥75  (GAA-FGF14 N=7; Austrian Study N=147) | 7 (100.0%) | 109 (74.1%) | 0.194 | 4 (57.1%) | 65 (44.2%) | 0.757 |

**Supplementary Table 3. Data and *p* values relative to the comparison between groups according to clinical variables.** Fisher’s exact test was used to assess differences between groups. *Legend*: SCP, superior cerebellar peduncles; WMA, white matter abnormalities; CC, corpus callosum; *, missing values for 4 subjects.

| ***Imaging feature*** | **Disease duration  ≤ 5 years (n=11)** | **Disease duration  > 5 years (n=24)** | **p value** | **Age at MRI  ≤ 60 (n=14)** | **Age at MRI > than 60 (n=21)** | **p value** | **Episodic ataxia at MRI*(n=2)** | **Permanent ataxia at MRI* (n=29)** | **p value** | **GAA size <300 (n=4)** | **GAA size ≥300 (n=31)** | **p value** |
| --- | --- | --- | --- | --- | --- | --- | --- | --- | --- | --- | --- | --- |
| Cerebral atrophy (n=15) | 3 | 12 | 0.075 | 3 | 12 | **0.046** | 0 | 14 | 0.488 | 2 | 13 | 1 |
| Brainstem atrophy (n=7) | 2 | 5 | 1 | 0 | 7 | **0.02** | 0 | 7 | 1 | 1 | 6 | 1 |
| Total cerebellar atrophy (n=33) | 10 | 23 | 0.53 | 14 | 19 | 0.77 | 2 | 28 | 1 | 3 | 30 | 0.218 |
| Isolated vermian atrophy (n=11) | 5 | 6 | 0.263 | 9 | 2 | **0.023** | 2 | 8 | 0.098 | 2 | 9 | 0.575 |
| SCP involvement (n=22) | 7 | 15 | 1 | 14 | 8 | **0.0002** | 2 | 17 | 0.509 | 2 | 20 | 0.617 |
| periventricular WMA (n=28) | 9 | 19 | 1 | 10 | 18 | 0.4 | 2 | 23 | 1 | 4 | 24 | 0.562 |
| multifocal WMA (n=22) | 6 | 16 | 0.707 | 8 | 14 | 0.72 | 1 | 21 | 0.503 | 3 | 19 | 1 |
| CC thinning (n=13) | 4 | 9 | 1 | 2 | 11 | **0.033** | 0 | 12 | 0.509 | 2 | 11 | 0.618 |
| ventricular enlargement (n=13) | 2 | 11 | 0.15 | 2 | 11 | **0.033** | 0 | 13 | 0.496 | 3 | 10 | 0.134 |
